# Social media and smartphone app use predicts maintenance of physical activity during Covid-19 enforced isolation in psychiatric outpatients

**DOI:** 10.1101/2020.06.26.20141150

**Authors:** Agnes Norbury, Shelley H Liu, Juan José Campaña-Montes, Lorena Romero-Medrano, Maria L. Barrigon, Emma Smith, MEmind Study Group, Antonio Artes, Enrique Baca-Garcia, M. Mercedes Perez-Rodriguez

**Affiliations:** Department of Psychiatry, Icahn School of Medicine at Mount Sinai, New York, NY, USA; Department of Population Health Science and Policy, Icahn School of Medicine at Mount Sinai, New York, NY, USA; Evidence-Based Behavior, Spain; Department of Signal Theory and Communications, Universidad Carlos III de Madrid, Spain; Department of Psychiatry, University Hospital Jimenez Diaz Foundation, Madrid, Spain; Instituto de Investigaciones Sanitarias Gregorio Marañón, Madrid, Spain; CIBERSAM, Carlos III Institute of Health, Madrid, Spain; Department of Psychiatry, Madrid Autonomous University, Madrid, Spain; Department of Psychiatry, University Hospital Rey Juan Carlos, Mostoles, Spain; Department of Psychiatry, General Hospital of Villalba, Madrid, Spain; Department of Psychiatry, University Hospital Infanta Elena, Valdemoro, Spain; Universidad Catolica del Maule, Talca, Chile; Department of Psychiatry, Centre Hospitalier Universitaire de Nîmes

## Abstract

There is growing concern that the social and physical distancing measures implemented in response to the Covid-19 pandemic may negatively impact health in other areas, via both decreased physical activity and increased social isolation. Here, we investigated whether increased engagement with digital social tools may help mitigate effects of enforced isolation on physical activity and mood, in a naturalistic study of at-risk individuals. Passively sensed smartphone app use and actigraphy data were collected from a sample of psychiatric outpatients before and during imposition of strict Covid-19 lockdown measures. Data were analysed using Gaussian graphical models: a form of network analysis which gives insight into the predictive relationships between measures across timepoints. Within-individuals, we found evidence of a positive predictive path between digital social engagement, general smartphone use, and physical activity – selectively under lockdown conditions (*N*=127 individual users, *M*=6201 daily observations). Further, we observed a positive relationship between social media use and total daily steps across individuals during (but not prior to) lockdown. Although there are important limitations on the validity of drawing causal conclusions from observational data, a plausible explanation for our findings is that, during lockdown, individuals use their smartphones to access social support, which may help guard against negative effects of in-person social deprivation and other pandemic-related stress. Importantly, passive monitoring of smartphone app usage is low burden and non-intrusive. Given appropriate consent, this could help identify people who are failing to engage in usual patterns of digital social interaction, providing a route to early intervention.

## Introduction

The novel coronavirus (Covid-19) pandemic is a major public health emergency. Responding to this crisis has required the implementation of unprecedented social distancing measures, including non-essential business closures, travel restrictions, and lockdown orders (*1*). As lockdowns become prolonged, researchers have drawn attention to potential negative effects of these measures, particularly in vulnerable populations (*2, 3*). These potential negative effects are twofold: decreases in physical activity lead to increased risk for cardiovascular disorders and other chronic health conditions (*4, 5*), whilst social isolation is linked to increased mortality and poorer mental health (*6*).

Although social media and smartphone use have been proposed to have negative effects on mood and mental health, the strength and reliability of evidence for this assertion is disputed, and the reality is likely to be more nuanced (*7, 8*). In particular, it has been suggested that digital social tools both help build community among people from historically marginalized populations (*9*), and enable individuals to foster and maintain social support networks during periods of stress and isolation (*10*). Differences in use of online social support resources may therefore significantly influence the extent to which people experience ‘social isolation’ during enforced physical distancing (*11*). Further, the ability to engage such compensatory mechanisms may be particularly important for individuals likely to be more vulnerable to the effects of enforced isolation – including people with pre-existing psychological distress (*12*), and other chronic health conditions that may increase risk of poor Covid-19 outcomes (*13*).

Here, we tested the hypothesis that people who engaged in more digital social interaction would be less vulnerable to negative effects of Covid-19 social distancing measures. Specifically, we investigated whether increased time spent using social media apps would predict maintenance of higher physical activity levels, pre-*vs* post-imposition of lockdown conditions. To address this question, we analysed passively sensed app use and physical activity (step count) data, collected from a sample of 163 psychiatric outpatients in Madrid, Spain. Data were available both before and during Covid-19 lockdown in Spain – to date one of the world’s countries most strongly affected, in terms of measurable impact on physical activity (*14, 15*). In a subset of participants (*N*=54 users), ecological momentary assessment of self-reported emotional state data were also available. This information was used to explore the idea that increased social media use may help protect against negative effects of lockdown-induced isolation on mood – either directly, or indirectly, via increased physical activity.

Data were analysed using a form of vector autoregression, where relationships between measures are represented as Gaussian graphical models (*16, 17*). This dynamic network-based approach has previously been successfully applied to temporally-ordered data in order to examine which of a set of inter-related variables predict each other at successive timepoints (*18–20*) – giving insight into Granger-causal relationships between measures (*21*). Although we note important limits on ability to draw causal inferences from observational data, in particular that there are likely to be unmeasured time-varying confounding factors that may result in statistical dependency between observed variables (*22*), this method of analysis can be used to highlight potential intervention points for users who fail to show adaptive behaviour patterns, and may therefore be at risk of poorer future outcomes (*3*).

## Results

### Participants

To avoid differences in sample characteristics across models, only users who had enough data to contribute to both pre- and post-lockdown network estimates were included in each analysis. Subsequently, data from *N*=127 users (92 female) were used to estimate physical activity and smartphone use networks (**Table 1**). In this sample, the most commonly represented psychiatric diagnosis was an anxiety, trauma, or stress-related disorder (56%), followed by unipolar or bipolar depression (40%). 22% of participants also had a diagnosis of a medical condition that would put them at increased risk from Covid-19 infection (*13*).

**Table 1.**
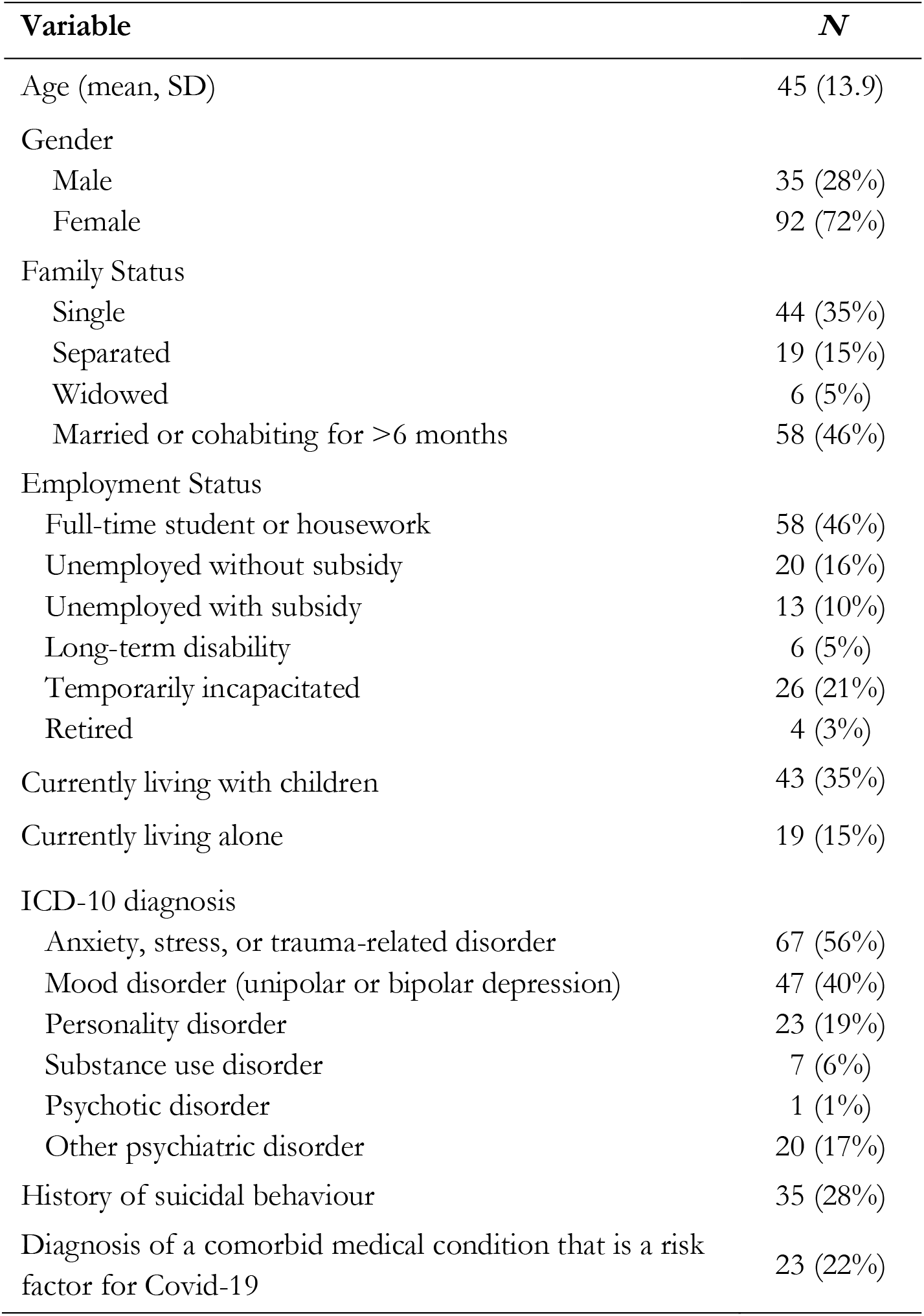
Demographic and clinical information for the study sample (*N*=127). Data represent *N* and percentage of available data, unless otherwise specified. *N*=7 (5.5%) participants were missing information about psychiatric diagnoses; *N*=21 (16.5%) participants were missing information about medical comorbidities. *ICD-10*, International Classification of Diseases, 10^th^ Edition. Psychiatric diagnosis categories are non mutually-exclusive. History of suicidal behaviour was defined as at least one suicide attempt or emergency room visit as a result of suicidal ideation. Comorbid medical conditions that were considered to place individuals at increased risk from Covid-19 were chronic pulmonary disease, chronic liver or kidney disease, cardiovascular disease, diabetes, hypertension, immunosuppressive disorder, clinical obesity, or cancer.

| Variable | N |
| --- | --- |
| Age (mean, SD) | 45 (13.9) |
| Gender |  |
| Male | 35 (28%) |
| Female | 92 (72%) |
| Family Status |  |
| Single | 44 (35%) |
| Separated | 19 (15%) |
| Widowed | 6 (5%) |
| Married or cohabiting for >6 months | 58 (46%) |
| Employment Status |  |
| Full-time student or housework | 58 (46%) |
| Unemployed without subsidy | 20 (16%) |
| Unemployed with subsidy | 13 (10%) |
| Long-term disability | 6 (5%) |
| Temporarily incapacitated | 26 (21%) |
| Retired | 4 (3%) |
| Currently living with children | 43 (35%) |
| Currently living alone | 19 (15%) |
| ICD-10 diagnosis |  |
| Anxiety, stress, or trauma-related disorder | 67 (56%) |
| Mood disorder (unipolar or bipolar depression) | 47 (40%) |
| Personality disorder | 23 (19%) |
| Substance use disorder | 7 (6%) |
| Psychotic disorder | 1 (1%) |
| Other psychiatric disorder | 20 (17%) |
| History of suicidal behaviour | 35 (28%) |
| Diagnosis of a comorbid medical condition that is a risk factor for Covid-19 | 23 (22%) |

As emotion self-reporting was entirely voluntary, mood data were only available in a subset of participants (*N*=54). Following restriction of users to individuals with enough data to be included in both models, *N*=22 users (14 female) provided enough emotional state information to be included in the pre- and post-lockdown networks for physical activity, smartphone use, and mood (**Table S2**). This reduction in sample size is a result of the fact that many users stopped providing emotion ratings during lockdown. Exploratory follow-up analysis revealed no significant differences in clinical/demographic features between the two samples (no difference in gender balance, family or employment status, proportion with an anxiety, mood, personality disorder or covid-19 risk diagnosis, or proportion with history of suicidal behaviour; *p*>0.26, Pearson’s Chi-squared tests; or age; *p*=0.49, one-way ANOVA), although the difference in group size renders formal comparison somewhat problematic. Further, as post-lockdown dropout in self-reporting of emotions is unlikely to be at random (e.g., may represent users most psychologically affected by the pandemic), we consider that users who continued to provide emotion data during lockdown may be unrepresentative of the sample as a whole, and results of the mood analysis should be interpreted with caution.

### Effect of lockdown on the relationship between physical activity and social media use

The population-level effects of lockdown and associated social distancing measures on daily activity in this cohort are explored in detail elsewhere (*23*). As reported previously, lockdown had a clear effect on daily physical activity (decreased step count) and social media usage (increased time spent using social apps) (**Figure 1a)**. Here, we focus on how within-user changes in social media use were related to future physical activity (step count), and whether the relationship between these measures was altered during enforced social isolation (lockdown).

**Figure 1.**
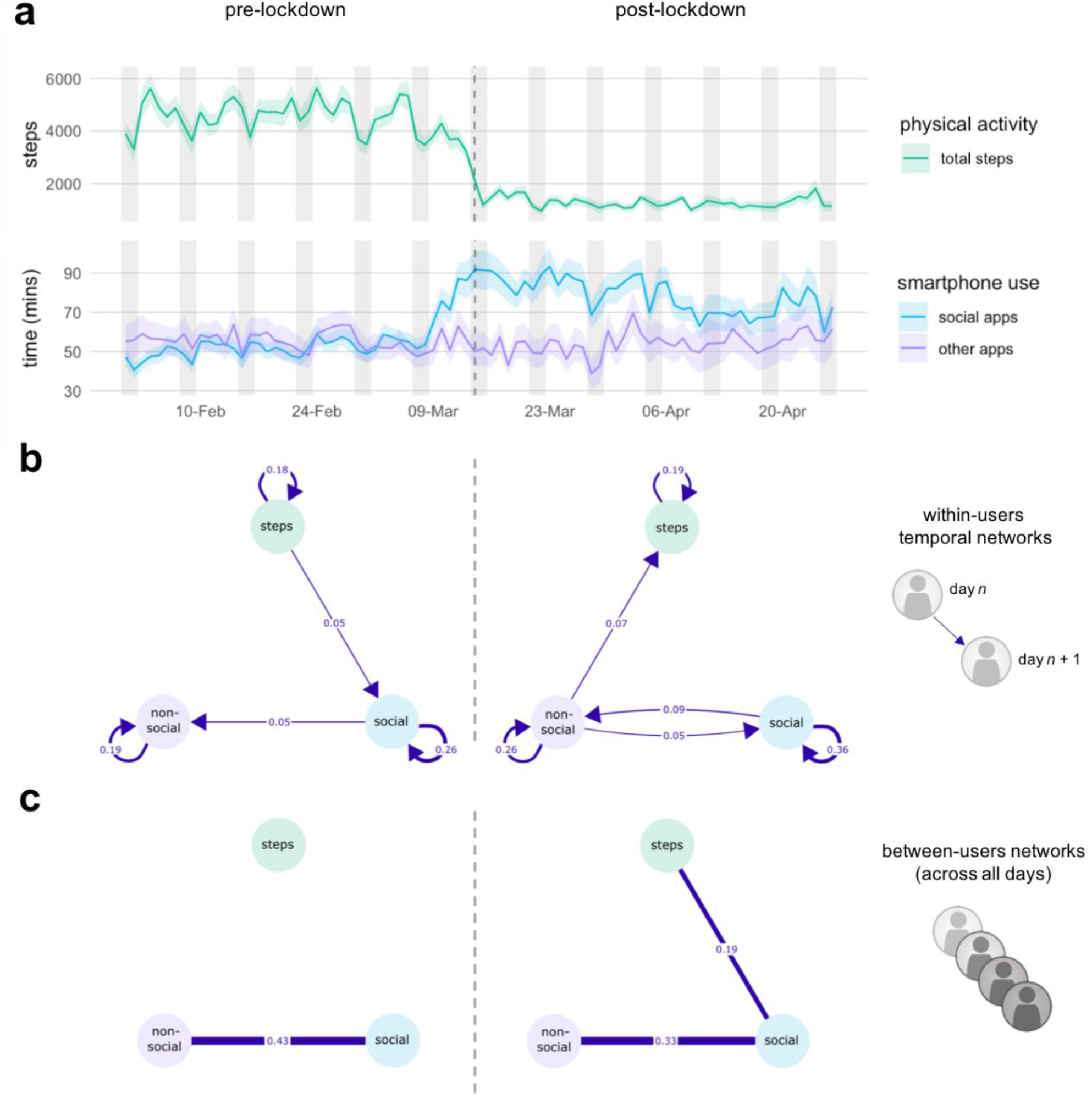
Effect of Covid-19 lockdown on the relationship between physical activity and social media use. **a** Mean (SE) of daily physical activity (step count), social, and non-social app use, as measured by passive smartphone sensing (eB^2^ monitoring app). The vertical dotted line represents the declaration of a national emergency (and associated lockdown measures) in Spain on 14/03/20. Vertical shading represents weekends (Saturday and Sunday). **b** Within-user temporal networks, pre- and post- imposition of lockdown conditions. The same *N*=127 users were included in each model. The pre-lockdown model included 38 time points (days), and the post-lockdown model included 45 time points. Blue lines represent positive predictive values for a given variable on day *n* on the value of the connected variable on day *n*+1, in the direction indicated by the arrowhead. Edges (connections between nodes) that do not significantly differ from 0 at alpha=0.05 are not depicted. **c** Between-users networks, representing covariance of means across participants, pre- and post-lockdown (derived from the same data as **b**).

To address this question, passively sensed smartphone use and actigraphy data were analysed using multilevel vector autoregression (VAR) (*17*). A key output of this analysis are Gaussian graphical models referred to as *temporal networks*: directed networks of regression coefficients that identify how well each variable predicts measures in the network across successive (lagged) time points (*16, 17*).

As there were clear effects of lockdown on daily activity levels, and VAR models assume stationarity (constancy of statistical properties over time), networks were estimated separately for the periods before and during lockdown implementation (*24*). Non-social (all other) app use was included in the models in order to account for potential nonspecific relationships between smartphone use and physical activity (as Gaussian graphical models are based on partial correlations, connections between nodes represent relationships that remain after adjusting for the values of all other variables in the network, (*25*)). Results of *between-subjects networks*, which represent the covariation of means across users, are also reported, in order to give insight into individual differences (*17*).

3280 observations over 38 time points (days) were included in the pre-lockdown analysis, and 2921 observations over 45 timepoints in the post-lockdown analysis (6201 total). Model comparison was used to compare networks with different degrees of temporal lagging between observations. For both time periods, this procedure favoured a 1-lag model (see Supplementary Material). The temporal networks presented here therefore represent whether deviations from an individual user’s mean for a particular measure on day *n* predict deviations in the value of other measures on day *n*+1.

#### Within-user temporal networks

Temporal networks depicting predictive relationships between changes in smartphone use and physical activity are shown in **Figure 1b**. In the pre-lockdown model, all three nodes had significant positive self-connections (auto-correlations) – indicating that an increase in number of steps taken, social media app use, or non-social smartphone app use on a given day predicted an increase in the same measure on the next day (fixed effect estimates [SE] of 0.176[0.026], 0.259[0.024], 0.190[0.028], respectively; *p*<0.001). Significant positive predictive paths also emerged from daily step count to social app use (fixed effect estimate 0.046[0.019], *p*=0.014), and from social app use to non-social app use (fixed effect estimate 0.051[0.023], *p*=0.030) – suggesting that increases in physical activity tended to predict increases in digital activity the next day (all other connections *p*>0.3; **Table S3**). Out-strength, a measure of which variables in the network are most strongly predictive of other network variables at the next time point, was equal for daily steps and social media use (both=0.055), and lower for non-social smartphone use (0.036).

In the post-lockdown model, positive self-connections for each node were still evident (fixed effect estimates [SE] of 0.195[0.028], 0.361[0.027], 0.259[0.025], *p*<0.001), but the direction of predictive effects *between* nodes appeared to have reversed. Specifically, the temporal network now revealed a positive predictive path from social to non-social app use (fixed effect estimate 0.088[0.026], *p*=0.001), and from non-social app use to daily step count (fixed estimate 0.067[0.021], *p*=0.001). There was also a mutually reinforcing loop from non-social back to social app use (fixed effect estimate 0.048[0.020], *p*=0.017; **Table S4**). This suggests that, during lockdown, increased social media use tended to result in increased next day physical activity, via a positively reinforcing loop that included greater general smartphone use. Subsequently, in the post-lockdown temporal network, out-strength was greater for digital than physical activity variables (social app use, 0.104; non-social app use 0.115; total steps, 0.031).

The case-drop bootstrap analysis revealed good stability of estimated temporal networks: in general, edges detected in the original analysis were included in the bootstrapped models at high frequencies, and edges absent in the original analysis were included at low frequencies (**Tables S5-6**; **Figure S3**). In particular, strong stability was observed for auto-correlations (included in 1000/1000 bootstrap models for both the pre- and post-lockdown networks). The predictive path between social and non-social app use demonstrated only moderate stability on the pre-lockdown model, but strong stability in the post-lockdown model (included in 448 and 966 bootstraps, respectively). Of note, the positive path between non-social app use and next-day physical activity was absent in all but 4/1000 bootstraps of the pre-lockdown network, but present in 921/1000 bootstraps of the post-lockdown network, indicating excellent stability for presence/absence of this path across time-periods. Simulation analysis revealed good to excellent power-related properties for fixed effects at *N*=127. Pre-lockdown, mean sensitivity across simulated datasets was 0.964, specificity was 0.878, and correlation between simulated and ‘true’ temporal network parameters was 0.984. Post-lockdown, mean sensitivity was 0.992, mean specificity was 0.777, and correlation was 0.992. Therefore, for both time periods, ability to detect ‘true’ present network connections (sensitivity) was excellent, with slightly reduced ability to detect ‘true’ absent connections (specificity).

#### Between-user networks

Between-users networks, representing the covariation of means across participants over all time points, provided further evidence for a relationship between social media use and physical activity levels, that emerged selectively during lockdown (**Figure 1c**). Users who, on average, spent more time engaging with social media apps, also, on average, took more daily steps – only in the *post*-lockdown network (post-lockdown correlation estimate=0.193, *p*=0.025; pre-lockdown correlation estimate=0.118, *p*=0.200; **Tables S3-4**). The fact that this relationship was evident only in the second model suggests that it is unlikely to be purely the result of an unmeasured covariate that would be common across time periods – such as a tendency for greater physical activity and greater smartphone use in younger participants.

The case-drop bootstrap analysis revealed that between-users relationship between social app use and daily step count was included in 298/1000 bootstraps of the pre-lockdown model, and 515/1000 bootstraps of the post-lockdown model: indicating moderate stability, with some evidence that the true pre-lockdown network may be less sparse than indicated here (i.e., that the between-subjects relationship between social app use and step count may be weaker, rather than absent, when not under lockdown conditions; **Table S5-6**; **Figure S3**).

### Effect of lockdown on the relationship between physical activity, social media use, and self-reported mood

Across the cohort who provided self-reported emotion data, mood tended to be negative (mean emotional valence rating <0; **Figure 2a**), but was more likely to be negative under lockdown (*23*). The pre-lockdown model included 324 observations over 36 time points (days), and the post-lockdown model included 122 observations over 34 time points (446 total observations). Model comparison again favoured a 1-lag model (see Supplementary Material).

**Figure 2.**
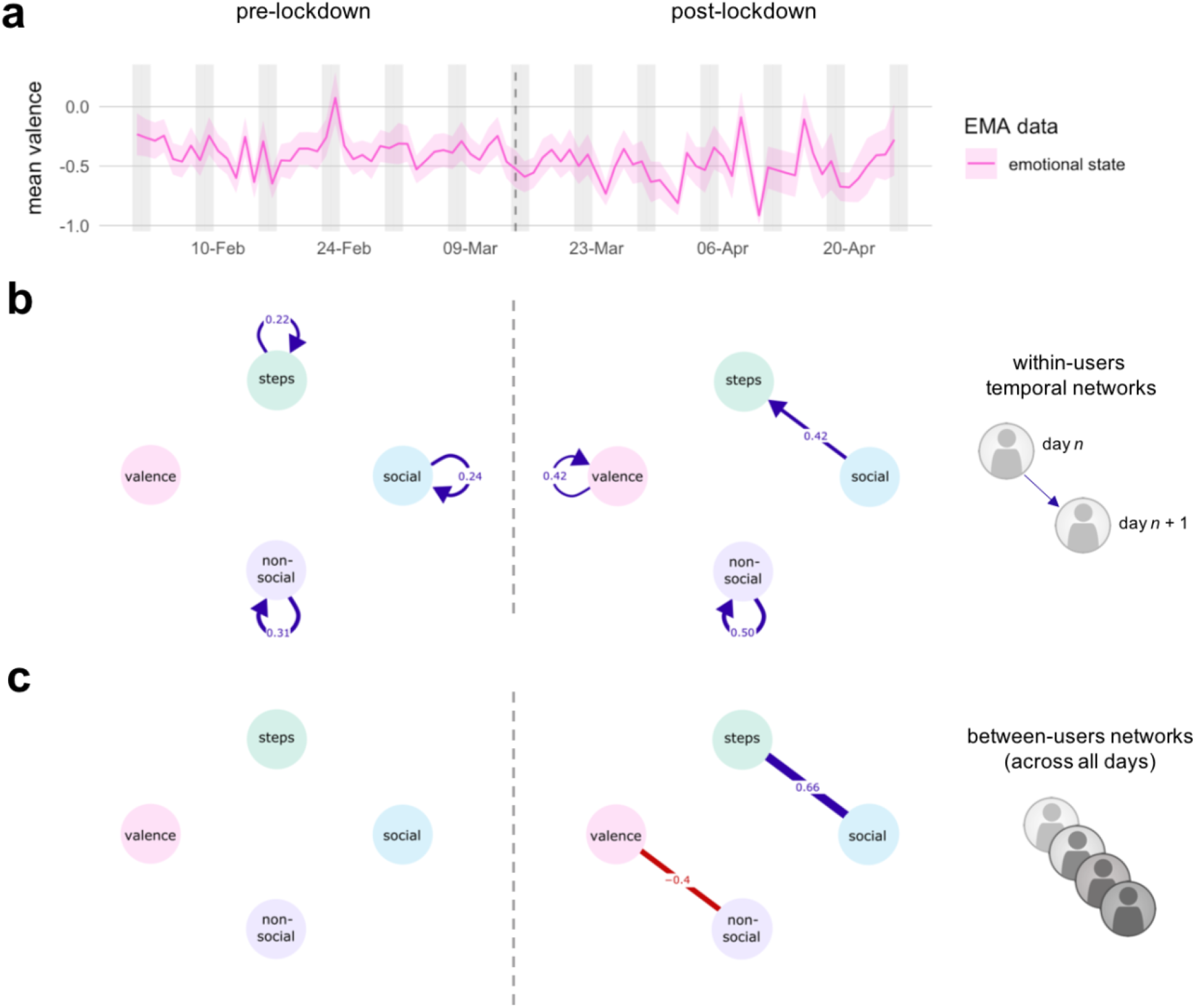
Effect of Covid-19 lockdown on the relationship between physical activity, social media use, and self-reported mood. **a** Mean (SE) of daily physical activity (step count), social, and non-social app use, as measured by passive smartphone sensing, and ecological momentary assessment (EMA) of emotional state data, as entered by users on an ad-hoc basis in the eB^2^ monitoring app. The vertical dotted line represents the declaration of a national emergency (and associated lockdown measures) in Spain on 14/03/20. Vertical shading represents weekends (Saturday and Sunday). **b** Within-user temporal networks, pre- and post- imposition of lockdown conditions. The same *N*=22 users were included in each model. The pre-lockdown model included 3280 observations over 38 time points (days), and the post-lockdown model included 2921 observations over 45 days.. Blue lines represent positive predictive values for a given variable on day *n* on the value of the connected variable on day *n*+1, in the direction indicated by the arrowhead. Edges (connections between nodes) that do not significantly differ from 0 at alpha=0.05 are not depicted. **c** Between-users networks, representing covariance of means across participants, pre- and post-lockdown (derived from the same data as **b**).

#### Within-user temporal networks

Temporal networks depicting predictive relationships between changes in physical activity, smartphone use, and self-reported mood are shown in **Figure 2b**. In the pre-lockdown model of users who provided emotion data, only positive auto-correlations between activity measures were evident (total daily steps, fixed effect estimate [SD] of 0.215[0.080], *p*=0.007; social app use, 0.241[0.059], *p*<0.001; non-social smartphone use,0.312[0.062], *p*<0.001; mean emotional valence, *p*>0.6). In the post-lockdown model, there was a strong positive predictive relationship between social media use and next day step count (fixed estimate 0.423[0.183], *p*=0.020), after adjusting for self-reported mean daily emotional valence. However, at this sample size, there was no strong evidence of increased daily physical activity (steps) predicting increases in mean emotional valence (mood) the next day (fixed effect estimate 0.182[0.124], *p*=0.124), or of a direct pathway between physical activity and mood (*p*>0.5). Full statistics for pre- and post-lockdown networks incorporating mood are available in **Tables S7-8**.

When accounting for individual differences in self-reported mood, the positive predictive path between social media use and next day step count was included in 36/999 successfully estimated case-drop bootstrap models in the pre-lockdown period, and 368/788 estimable models in the post-lockdown period, indicating moderate stability for presence/absence of this connection at this sample size (**Tables S9-10**; **Figure S4**). Simulation analysis at *N*=22 users for the pre-lockdown period revealed mean sensitivity of 0.997, mean specificity of 0.688, and correlation of 0.826 for temporal fixed effects. Post-lockdown values were 1.00, 0.591, and 0.777, respectively. Therefore, while this analysis may have adequate sensitivity (ability to discover true connections), it may be under-powered with respect to specificity (ability to detect absent connections).

#### Between-user networks

Between-user networks are depicted in **Figure 2c**. Similar to the results from the larger sample, after adjusting for covariance with self-reported mood, there was a significant between-subjects association between physical activity and social media use in the post-lockdown network only (post-lockdown correlation estimate in users who volunteered emotional state information 0.699, *p*<0.001; pre-lockdown estimate in the same individuals 0.281, *p*=0.146). There was also a negative relationship between mood and non-social smartphone use, such that users who tended to report more negative mood also tended to spend more time each day using non-social smartphone apps.

The between-subjects covariation between social media use and physical activity was included in 340/999 bootstraps of the pre-lockdown model, and 786/788 bootstraps of the post-lockdown model. There was also some evidence for a positive between-subjects edge between mood and physical activity (tendency towards higher self-reported mood in individuals with higher daily steps counts, or *vice versa*), which we may be underpowered to detect here (edge included in 595/999 pre-lockdown and 519/788 post-lockdown bootstraps; **Tables S9-10**; **Figure S4**).

## Discussion

The findings presented here have implications for gauging the value of social media and smartphone use to psychologically and medically vulnerable individuals during periods of enforced physical isolation. Within-individuals, we identified a specific pattern of behaviour during lockdown – whereby increased social media and smartphone use on a particular day predicted an increased number of steps the next day. The overall predictive effect of smartphone use variables in the temporal network (out-strength) was greater post- *vs* pre- lockdown, suggesting greater influence on physical activity under social distancing conditions. We also found evidence of a positive relationship between social media use and total daily steps *across* individuals during (but not prior to) lockdown, suggesting that – specifically during social isolation – users who, on average, spent more time using social media also, on average, took more daily steps.

One plausible explanation for these findings is that, during lockdown, individuals make use of digital tools to harness online social support structures (*26*), which may mitigate the negative effects of in-person social deprivation and other pandemic-related stress (*27, 28*). Specifically, we hypothesize that social interaction may promote engagement in physical activity by guarding against inertia and apathy associated with low mood (*29, 30*). Possible mechanisms for this effect include stress-reducing effects of information-sharing or co-rumination (talking through problems together with others) (*31, 32*), positive mood contagion (*33, 34*), and general mood-boosting effects of social interaction (*35, 36*). Although we found no strong evidence of a pathway between social media use, physical activity, and self-reported emotional valence in our emotion-reporting subsample, simulation evidence suggested we lacked power to determine absence of effects in this analysis, and a positive predictive association between physical activity and mood has been well-established in previous work (*35, 37*). Importantly, increased physical activity may activate a positive feedback loop, whereby greater activity improves mood and therefore further enhances motivation to continue exercising. Indeed, both full-sample networks contained strong positive self-loops for total daily steps, suggesting that physical activity on day *n* is robust predictor of activity on day *n*+1. However, it should be noted that the validity of interpreting these results in a causal way is limited by several key assumptions underlying our analysis. In particular, it is possible that some unmeasured factor (e.g., variation in psychological/somatic symptoms, or contextual factors such as weather) influenced both smartphone use and physical activity with varying time delays, resulting in spurious dependencies between the two (*22*). Further, although we attempted to correct for one form of seasonality in our time series data (weekday *vs* weekend effects), the assumption of stationarity across the 5-6 week time periods before/after lockdown might not be reasonable, which may limit our ability to differentiate within- and between-users effects (*38*). Finally, the methods employed here are exploratory, and should primarily be considered as hypothesis-generating, rather than confirmatory (*17*). Ultimately, a causal role for digital social interaction in promoting physical activity levels under different conditions will best be confirmed using data from interventional study designs.

An advantage of the data presented here is that it is truly prospective and longitudinal with respect to onset of the Covid-19 pandemic and associated social distancing measures. The measures from which our major conclusions are drawn are passively sensed – requiring no active data submission on behalf of the participant – which may help guard against sampling-related biases (e.g. collider bias, (*39, 40*)), whilst also decreasing measurement error (particularly compared to self-report, which may not be a very accurate measure of online activity, (*41*)). Further, the sample for the main analysis was representative of the target population (85-87% recruitment rate from the outpatient clinic in parent studies, (*42*)), and users were behaving naturalistically in the community, free from the influence of perceived experimenter demands. Although we found no evidence for differences in clinical or sociodemographic features between users included in the main analysis and mood sub-sample, the latter is likely to be affected by self-selection bias (see Results), therefore we focus our discussion on findings from the larger group.

Our study has several important limitations. Consistent with previous observations, the predictive (cross-lagged) associations we observed *between* measures were substantially smaller than autocorrelation effects (*18*). Although fixed effect estimates for temporal models reported here were modest, the relatively large standard deviations of the random effects estimates indicate that the strength of these effects may vary significantly across individuals (e.g. fixed effects estimate of 0.088 with random effects SD of 0.159 for social -> nonsocial app use; and fixed effect estimate of 0.067 with random effects SD of 0.079 for nonsocial app use -> step count; **Table S4**). Our sample size did not allow us to break down users by important between-subjects variables such as psychiatric diagnosis, age group, presence of a Covid-19 risk comorbidity, or household status, which might reasonably be expected to affect both baseline behaviour and moderate the impact of lockdown. If and how this pattern varies across these important between-subjects dimensions should be addressed in future research. Further, by focusing on total daily time as the unit of analysis for smartphone use, we may be masking more fine-grained effects of application type and usage patterns – including differential effects of active communication/information-gathering and more passive browsing styles on mood (*43–46*). Under our chosen analysis framework, effects that occur at faster timescale will be classified as contemporaneous effects, and effects which occur over substantially longer timescales may be incorporated into between-subjects variance (*38, 20*). Although in the current dataset model comparison favoured a simple 1-lag structure, it is possible that longer-term dependencies between variables might be revealed by a more complex multi-timescale analysis.

Previous research has highlighted the importance of maintaining physical activity during periods of isolation, in terms of both physical and mental wellbeing (*2, 4, 5*). This is likely to be particularly important for people with a diagnosis of a psychiatric disorder, as they are already significantly less likely to meet guideline physical activity levels (*47, 48*). Here, we present preliminary evidence of a positive predictive path between digital social engagement and physical activity, under conditions where physical activity levels are suppressed. Given users’ consent, monitoring of the relationship between social media and physical activity data could potentially help identify individuals who are failing to engage this mechanism, providing a route to early intervention in vulnerable populations (either via a clinician, or more informally, e.g. via smartphone prompts) (*3*). Successful translation of this approach to clinical practice will likely require consultation with patient groups and other stakeholders, who may hold concerns about secondary data use and other ethical issues related to the use passive monitoring technologies in psychiatry (*49, 50*). Finally, in addition to having implications for how best to maintain physical and mental health under lockdown conditions, these results contribute to a broader discussion on the role of digital technologies in wellbeing. We second recent calls for a greater emphasis on nuance in this debate, in particular by focusing on longitudinal assessments, and paying greater attention to both specific populations and contexts under which phenomena are observed (*9, 51*).

## Methods

### Data and code availability statement

Data are not freely publicly available due to lack of participant consent for public sharing, but are available from the authors upon request. To facilitate reproducibility, synthetic data generated using the R package synthpop (*52*) is available alongside analysis code and R package version information at https://github.com/agnesnorbury/app-use_physical-activity_covid19.

### Participants

Data were drawn from two ongoing studies of psychiatric outpatients in Madrid, Spain that involve remote smartphone monitoring (*53, 54*). Both studies received ethical approval from the Fundación Jimenez Diaz Hospital Institutional Review Board, and all participants provided written informed consent. Participants were required to be age 18 or older, fluent in Spanish, and to possess a smartphone with internet access. Sociodemographic and clinical information were collected via an electronic health record tool (MEmind, (*55*)). Analyses were restricted to users with 20 or more days of physical activity data, to enable stable network estimation (total *N*=163).

### Passively-sensed physical activity and smartphone use data

Physical activity (daily step count) and smartphone use (time each day spent using applications) were collected using the Evidence-Based Behaviour (eB^2^) monitoring app (*56*). Social media use was defined as time spent using apps from the ‘communication’ (e.g., WhatsApp, Facebook Messenger) or ‘social’ (e.g., Facebook, Instagram, TikTok) app store categories (*57, 58*). Non-social app use was defined as time spent using any other type of smartphone app (e.g., games, tools, travel apps, audiovisual media players).

### Self-reported emotional state data

Users were able to enter information about their current emotional state within the eB^2^ app, by selecting from a range of visual icons (**Figure S1**). Emotion self-reporting was voluntary and un-prompted, and therefore was only available for a subset of users and days. Recorded emotions were recoded as positive (happy, delighted, motivated, relaxed), neutral (neutral), or negative (angry, sad, fearful, disgusted, tired, in pain, worried/overwhelmed). Emotions from each category were scored +1, 0, or −1, respectively, and scores were averaged across each day to generate an index of mean daily emotional valence (“mood”).

### Network analysis of physical activity, smartphone use, and mood data

#### Data preprocessing

As physical activity and app use data exhibited significant weekly variation (lower values at weekends, see **Figure 1a**), these data were first adjusted for 7-day seasonality. Specifically, data were decomposed into weekly seasonal variation, overall trend, and residual time series components using the R function stl (*59*), following which the seasonality component was subtracted from the overall time series. Plots of the mean detrended times series data are available in **Figure S2**. Physical activity, app use, and emotional valence data were also observed to be significantly skewed, and so were transformed prior to network estimation in order to meet assumptions of the Gaussian graphical model (nonparanormal transformation as implemented in function npn from the R package huge, (*60*)). Results of this transformation on distributions of observations across users are depicted in **Table S1**.

#### Network estimation

Networks were estimated using two-step multilevel vector autoregression (*17*), as implemented in the R package mlVAR, version 0.4.4 (*61*), using a two-step frequentist estimation procedure (mlVAR estimation option lmer). This involves running sequential univariate multilevel regression models on previous measurements, with within-subject centred lagged variables as within-subjects level predictors, and the sample means of all other variables as between-subjects predictors (for full details see (*16, 17*)). This analysis yields three kinds of network: a *temporal network*, which consists of a directed network of regression coefficients between current and lagged variables; a *between-subjects network*, which describes relationships between the stationary means of subjects; and a *contemporaneous network*, which describes relationships within a single measurement occasion, after controlling for temporal effects (*17*). Here, we report findings from the first and second kind of networks, as we were primarily interested in how study variables predicted each other over time, and the extent to which this might vary across subjects. Findings for the third kind of network (contemporaneous) are reported in the Supplementary Material for completeness, but are not discussed further. As the number of variables in our networks was small, random effects in temporal networks were allowed to be correlated. Bayesian Information Criterion (BIC) scores were used to compare models incorporating different numbers of time lags, penalized for model complexity. Out-strength was calculated as the absolute temporal edge weights extending out from each node, excluding autocorrelations, as per (*18, 19*).

#### Stability and power assessment

Stability of network estimation was assessed using the case-bootstrap approach previously described by Epskamp and colleagues (*38, 62*). Specifically, 1000 bootstrapped models were estimated in which 25% of cases (users) were dropped at random, and the proportion of times each network connection (edge) was included in the bootstrap sample counted. Power of our analysis to detect temporal fixed-effects was further assessed via simulation, as per (*25, 38*). Specifically, simulated datasets (100 per condition) were generated using the estimated (‘true’) model parameters, at different sample sizes. Sensitivity (rate of discovery of true network connections), specificity (rate of discovery of absent network connections), and correlation between networks estimated from the simulated and ‘true’ datasets are then reported.

#### Visualisation

Networks are depicted using the colourblind-friendly theme included in mlVAR: with positive edges coloured blue, and negative edges coloured red. For clarity, edges that did not significantly differ from 0 at alpha=0.05 are not plotted. For the between-subjects networks, edges (connections between nodes) were considered significant according to an ‘and’ rule (i.e., only retained if both edges on which a connection was based were significant at alpha=0.05). Edges are drawn with line widths proportional to effect size.

## Data Availability

Data associated with this publication is not freely publicly available due to lack of participant consent for public sharing, but is available upon from the authors upon request. Code used to generate all results described here is available at https://github.com/agnesnorbury/app-use_physical-activity_covid19. Code is accompanied by synthetic data generated using the R package synthpop, in order to facilitate analytic reproducibility.

## Notes

### Competing Interest Statement

Dr. Perez-Rodriguez has received research grant funding from Neurocrine Biosciences (Inc), Millennium Pharmaceuticals, Takeda, Merck, and AI Cure; She is an Advisory Board member for Neurocrine Biosciences, Inc, and a consultant on an American Foundation for Suicide Prevention (AFSP) grant (LSRG-1-005-16, PI: Baca-Garcia).
Enrique Baca-Garcia has been a consultant to or has received honoraria or grants from Janssen Cilag, Lundbeck, Otsuka, Pziffer, Servier and Sanoffi.
Antonio Artes, Juan José Campaña-Montes and Enrique Baca-Garcia are founders of eB2. Enrique Baca-Garcia has designed MEmind.

### Funding Statement

Partly funded by Carlos III (ISCIII PI16/01852), American Foundation for Suicide Prevention (LSRG-1-005-16), the Madrid Regional Government (B2017/BMD-3740 AGES-CM 2CM; Y2018/TCS-4705 PRACTICO-CM), MINECO/FEDER ('ADVENTURE', id. TEC2015-69868-C2-1-R), MCIU Explora Grant aMBITION' ('id. TEC2017-92552-EXP) and MICINN ('CLARA', id. RTI2018-099655-B-I00). Funding sources had no role in the design or conduction of the study.

### Author Declarations

Institutional Review Board at the Psychiatry Department of Fundación Jimenez Diaz Hospital

