## Supplementary Material for "Social media and smartphone app use predicts maintenance of physical activity during Covid-19 enforced isolation in psychiatric outpatients"

**Supplementary Methods**
All statistical analyses were carried out in R, version 3.6.1 (R Core Team, 2019). Version information for packages on which the analysis depended (R sessionInfo output) is available at <https://github.com/agnesnorbury/app-use_physical-activity_covid19>.

Following informed consent, the eB^2^ app was installed on participants’ smartphones and configured with the assistance of a physician. After this time, the application passively collected smartphone use and actigraphy data, without requiring further input from the user (users were able to uninstall the app at any point).

**Supplementary Results**

Model comparison was used to compare networks with different degrees of temporal lagging between observations. For both time periods this procedure favoured a 1-lag model (pre-lockdown, mean Bayesian Information Criterion [BIC] across measures of 4976, compared to 5094 for a 1,2-lag model and 5269 for a 1,2,3-lag model; post-lockdown, mean BIC of 5141, compared to 5229 for a 1,2-lag model and 5417 for a 1,2,3-lag model).

For the mood sub-analysis, Model comparison again favoured a 1-lag model (pre-lockdown, mean BIC across measures for 1-lag model 687, compared to 843 for a 1,2-lag model; higher lag models and >1lag models for the post-lockdown time period were inestimable due to too few observations).

| **variable**  **(raw data)** | **values** | **histogram** |
| --- | --- | --- |
| nonsocial_usage | mean (SD) : 3294.6 (5047.1)  min < med < max: 0 < 1622 < 39007  IQR (CV) : 3059 (1.5) | 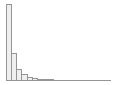 |
| social_usage | mean (SD) : 3982.3 (4169.5)  min < med < max: 1 < 2789 < 40000  IQR (CV) : 3956 (1) | 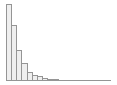 |
| steps_total | mean (SD) : 2883.4 (3896)  min < med < max: 0 < 1306 < 50000  IQR (CV) : 4028 (1.4) | 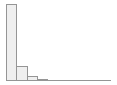 |

| **variable**  **(seasonality adjusted)** | **values** | **histogram** |
| --- | --- | --- |
| nonsocial_usage | mean (SD) : 3395.5 (4976.8)  min < med < max: -6302.5 < 1823.5 < 41484.7  IQR (CV) : 3047.4 (1.5) | 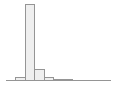 |
| social_usage | mean (SD) : 4082.4 (3977.3)  min < med < max: -5903.5 < 2974.5 < 43668.4  IQR (CV) : 3858.1 (1) | 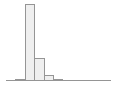 |
| steps_total | mean (SD) : 3006.7 (3691.4)  min < med < max: -6047.6 < 1784.1 < 48854.4  IQR (CV) : 4107.9 (1.2) | 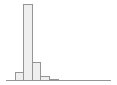 |

| **variable**  **(seasonality adjusted and npn transformed)** | **values** | **histogram** |
| --- | --- | --- |
| nonsocial_usage | mean (SD) : 0 (1)  min < med < max: -3.7 < 0 < 3.7  IQR (CV) : 1.3 (171180499863974752) | 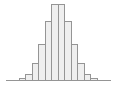 |
| social_usage | mean (SD) : 0 (1)  min < med < max: -3.7 < 0 < 3.7  IQR (CV) : 1.3 (168302943547881888) | 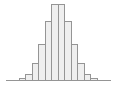 |
| steps_total | mean (SD) : 0 (1)  min < med < max: -3.7 < 0 < 3.7  IQR (CV) : 1.3 (211267341144909664) | 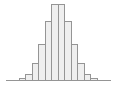 |

**Table S1**. **Effects of 7-day seasonality adjustment and nonparametric normal (npn) transformation on distribution of physical activity and smartphone use measures across users**. Tables generated with R package summarytools. *steps_total*, total daily step count; *social_usage*, total time spent using social media apps; *nonsocial_usage*, total time spent using any other app, per day. *IQR*, interquartile range; *CV*, coefficient of variation (relative standard deviation)

| **Variable** | ***N*** |
| --- | --- |
| Age (mean, SD) | 47 (12.0) |
| Gender  Male  Female | 8 (36%)  14 (64%) |
| Family Status  Single  Separated  Widowed  Married or cohabiting for >6 months | 10 (46%)  1 (5%)  1 (5%)  10 (46%) |
| Employment Status  Full-time student or housework  Unemployed without subsidy  Unemployed with subsidy  Long-term disability  Short-term disability  Retired | 10 (46%)  1 (5%)  3 (14%)  2 (9%)  6 (27%)  0 (0%) |
| Currently living with children | 6 (27%) |
| Currently living alone | 6 (27%) |
| ICD-10 diagnosis  Anxiety, stress, or trauma-related disorder  Mood disorder (unipolar or bipolar depression)  Personality disorder  Substance use disorder  Psychotic disorder  Other psychiatric disorder | 10 (50%)  10 (50%)  4 (20%)  1 (5%)  0 (0%)  4 (20%) |
| History of suicidal behaviour | 8 (36%) |
| Diagnosis of a comorbid medical condition that is a risk factor for Covid-19 | 2 (11%) |

**Table S2.** **Demographic and clinical information for the sub-sample of participants who provided emotional state data (*N*=22).** Data represent *N* and percentage of available data, unless otherwise specified. *N*=2 (9%) participants were missing information about psychiatric diagnoses; *N*=3 (14%) participants were missing information about medical comorbidities. *ICD-10*, International Classification of Diseases, 10^th^ Edition. Psychiatric diagnosis categories are non mutually-exclusive. History of suicidal behaviour was defined as at least one suicide attempt or emergency room visit as a result of suicidal ideation. Comorbid medical conditions that were considered to place individuals at increased risk from Covid-19 were chronic pulmonary disease, chronic liver or kidney disease, cardiovascular disease, diabetes, hypertension, immunosuppressive disorder, clinical obesity, or cancer.

Within-user temporal network:

| from | to | lag | fixed effect estimate | SE | *p* | random  effects SD |
| --- | --- | --- | --- | --- | --- | --- |
| steps_total | steps_total | 1 | 0.176 | 0.026 | 0.000 | 0.167 |
| steps_total | social_usage | 1 | 0.047 | 0.019 | 0.014 | 0.074 |
| steps_total | nonsocial_usage | 1 | 0.007 | 0.021 | 0.738 | 0.116 |
| social_usage | steps_total | 1 | -0.004 | 0.021 | 0.837 | 0.048 |
| social_usage | social_usage | 1 | 0.259 | 0.024 | 0.000 | 0.133 |
| social_usage | nonsocial_usage | 1 | 0.051 | 0.023 | 0.030 | 0.130 |
| nonsocial_usage | steps_total | 1 | 0.016 | 0.022 | 0.460 | 0.046 |
| nonsocial_usage | social_usage | 1 | 0.020 | 0.022 | 0.361 | 0.060 |
| nonsocial_usage | nonsocial_usage | 1 | 0.190 | 0.028 | 0.000 | 0.180 |

Between-users network:

| variable 1 | variable 2 | *p* 1->2 | *p* 1<-2 | pcor | cor |
| --- | --- | --- | --- | --- | --- |
| social_usage | steps_total | 0.200 | 0.048 | 0.141 | 0.118 |
| nonsocial_usage | steps_total | 0.443 | 0.281 | -0.081 | -0.023 |
| nonsocial_usage | social_usage | 0.000 | 0.000 | 0.428 | 0.422 |

Within-user contemporaneous network (estimated post-hoc):

| variable 1 | variable 2 | *p* 1->2 | *p* 1<-2 | pcor | cor |
| --- | --- | --- | --- | --- | --- |
| social_usage | steps_total | 0.239 | 0.325 | 0.024 | 0.029 |
| nonsocial_usage | steps_total | 0.516 | 0.386 | 0.016 | 0.023 |
| nonsocial_usage | social_usage | 0.000 | 0.000 | 0.249 | 0.250 |

**Table S3**. **Summary of pre-lockdown network analysis relating daily physical activity to smartphone use.** *N*=127 participants and 38 time points (days) were included in the model. The temporal network included a 1-timepoint (day) lag for predictive relationships. *steps_total*, total daily step count; *social_usage*, total time spent using social media apps; *nonsocial_usage*, total time spent using any other app, per day; *SE*, standard error; *SD*, standard deviation; *pcor*, partial correlation*; cor*, correlation.

Within-user temporal network:

| from | to | lag | fixed effect estimate | SE | *p* | random effects SD16 |
| --- | --- | --- | --- | --- | --- | --- |
| steps_total | steps_total | 1 | 0.195 | 0.028 | 0.000 | 0.216 |
| steps_total | social_usage | 1 | -0.006 | 0.015 | 0.684 | 0.049 |
| steps_total | nonsocial_usage | 1 | -0.025 | 0.018 | 0.169 | 0.079 |
| social_usage | steps_total | 1 | -0.016 | 0.021 | 0.454 | 0.070 |
| social_usage | social_usage | 1 | 0.361 | 0.027 | 0.000 | 0.191 |
| social_usage | nonsocial_usage | 1 | 0.088 | 0.026 | 0.001 | 0.159 |
| nonsocial_usage | steps_total | 1 | 0.067 | 0.021 | 0.001 | 0.079 |
| nonsocial_usage | social_usage | 1 | 0.048 | 0.020 | 0.017 | 0.096 |
| nonsocial_usage | nonsocial_usage | 1 | 0.259 | 0.025 | 0.000 | 0.155 |

Between-users network:

| variable 1 | variable 2 | *p* 1->2 | *p* 1<-2 | pcor | cor |
| --- | --- | --- | --- | --- | --- |
| social_usage | steps_total | 0.025 | 0.023 | 0.193 | 0.192 |
| nonsocial_usage | steps_total | 0.640 | 0.696 | -0.037 | 0.029 |
| nonsocial_usage | social_usage | 0.000 | 0.000 | 0.333 | 0.332 |

Within-user contemporaneous network (estimated post-hoc):

| variable 1 | variable 2 | *p* 1->2 | *p* 1<-2 | pcor | cor |
| --- | --- | --- | --- | --- | --- |
| social_usage | steps_total | 0.305 | 0.646 | 0.016 | 0.013 |
| nonsocial_usage | steps_total | 0.485 | 0.475 | -0.015 | -0.011 |
| nonsocial_usage | social_usage | 0.000 | 0.000 | 0.272 | 0.272 |

**Table S4**. **Summary of post-lockdown network analysis relating daily physical activity to smartphone use.** *N*=127 participants and 45 time points (days) were included in the model. The temporal network included a 1-timepoint (day) lag for predictive relationships. *steps_total*, total daily step count; *social_usage*, total time spent using social media apps; *nonsocial_usage*, total time spent using any other app, per day; *SE*, standard error; *SD*, standard deviation; *pcor*, partial correlation*; cor*, correlation.

| Number of times temporal effects were included | | | |
| --- | --- | --- | --- |
|  | steps_total | social_usage | nonsocial_usage |
| steps_total | **1000** | **573** | 3 |
| social_usage | 1 | **1000** | **448** |
| nonsocial_usage | 4 | 16 | **1000** |
| Number of times between-subjects effects were included | | | |
|  | steps_total | social_usage | nonsocial_usage |
| steps_total | - |  |  |
| social_usage | 298 | - |  |
| nonsocial_usage | 24 | **1000** | - |
| Number of times contemporaneous effects were included | | | |
|  | steps_total | social_usage | nonsocial_usage |
| steps_total | - |  |  |
| social_usage | 37 | - |  |
| nonsocial_usage | 18 | **1000** | - |

**Table S5. Stability of pre-lockdown network analysis relating daily physical activity to smartphone use**. Values represent number of times (out of 1000) each network connection was included in the 25% case-drop bootstrap models. Bold type face indicates effects which were included in the original analysis.

| Number of times temporal effects were included | | | |
| --- | --- | --- | --- |
|  | steps_total | social_usage | nonsocial_usage |
| steps_total | **1000** | 1 | 84 |
| social_usage | 1 | **1000** | **966** |
| nonsocial_usage | **921** | **581** | **1000** |
| Number of times between-subjects effects were included | | | |
|  | steps_total | social_usage | nonsocial_usage |
| steps_total | - |  |  |
| social_usage | **515** | - |  |
| nonsocial_usage | 1 | **1000** | - |
| Number of times contemporaneous effects were included | | | |
|  | steps_total | social_usage | nonsocial_usage |
| steps_total | - |  |  |
| social_usage | 20 | - |  |
| nonsocial_usage | 4 | **1000** | - |

**Table S6. Stability of post-lockdown network analysis relating daily physical activity to smartphone use**. Values represent number of times (out of 1000) each network connection was included in the 25% case-drop bootstrap models. Bold type face indicates effects which were included in the original analysis.

Within-user temporal network:

| from | to | lag | fixed effect estimate | SE | *p* | random  effects SD |
| --- | --- | --- | --- | --- | --- | --- |
| steps_total | steps_total | 1 | 0.215 | 0.080 | 0.007 | 0.218 |
| steps_total | social_usage | 1 | 0.021 | 0.051 | 0.675 | 0.122 |
| steps_total | nonsocial_usage | 1 | -0.059 | 0.037 | 0.111 | 0.030 |
| steps_total | valence | 1 | 0.004 | 0.061 | 0.942 | 0.166 |
| social_usage | steps_total | 1 | -0.120 | 0.087 | 0.166 | 0.176 |
| social_usage | social_usage | 1 | 0.241 | 0.059 | 0.000 | 0.086 |
| social_usage | nonsocial_usage | 1 | 0.014 | 0.079 | 0.858 | 0.217 |
| social_usage | valence | 1 | -0.064 | 0.073 | 0.385 | 0.159 |
| nonsocial_usage | steps_total | 1 | 0.071 | 0.105 | 0.495 | 0.267 |
| nonsocial_usage | social_usage | 1 | 0.014 | 0.061 | 0.824 | 0.087 |
| nonsocial_usage | nonsocial_usage | 1 | 0.312 | 0.062 | 0.000 | 0.101 |
| nonsocial_usage | valence | 1 | 0.155 | 0.105 | 0.141 | 0.331 |
| valence | steps_total | 1 | -0.076 | 0.083 | 0.358 | 0.156 |
| valence | social_usage | 1 | -0.087 | 0.050 | 0.082 | 0.018 |
| valence | nonsocial_usage | 1 | -0.079 | 0.055 | 0.149 | 0.085 |
| valence | valence | 1 | -0.035 | 0.068 | 0.610 | 0.172 |

Between-users network:

| variable 1 | variable 2 | *p* 1->2 | *p* 1<-2 | pcor | cor |
| --- | --- | --- | --- | --- | --- |
| social_usage | steps_total | 0.146 | 0.416 | 0.242 | 0.281 |
| nonsocial_usage | steps_total | 0.061 | 0.043 | 0.390 | 0.311 |
| nonsocial_usage | social_usage | 0.759 | 0.691 | -0.008 | 0.070 |
| valence | steps_total | 0.098 | 0.009 | 0.373 | 0.301 |
| valence | social_usage | 0.721 | 0.980 | 0.031 | 0.118 |
| valence | nonsocial_usage | 0.272 | 0.011 | -0.330 | -0.205 |

Within-user contemporaneous network (estimated post-hoc):

| variable 1 | variable 2 | *p* 1->2 | *p* 1<-2 | pcor | cor |
| --- | --- | --- | --- | --- | --- |
| social_usage | steps_total | 0.791 | 0.825 | 0.004 | -0.009 |
| nonsocial_usage | steps_total | 0.264 | 0.164 | -0.076 | -0.075 |
| nonsocial_usage | social_usage | 0.027 | 0.024 | 0.166 | 0.166 |
| valence | steps_total | 0.146 | 0.078 | 0.088 | 0.086 |
| valence | social_usage | 0.899 | 0.974 | -0.005 | -0.002 |
| valence | nonsocial_usage | 0.661 | 0.644 | 0.025 | 0.018 |

**Table S7**. **Summary of pre-lockdown network analysis relating daily physical activity to smartphone use and self-reported mood.** *N*=22 participants and 36 time points (days) were included in the model. The temporal network included a 1-timepoint (day) lag for predictive relationships. *steps_total*, total daily step count; *social_usage*, total time spent using social media apps; *nonsocial_usage*, total time spent using any other app, per day; *valence,* mean valence of emotions recorded each day (mood); *SE*, standard error; *SD*, standard deviation; *pcor*, partial correlation*; cor*, correlation.

Within-user temporal network:

| from | to | lag | fixed effect estimate | SE | *p* | random  effects SD |
| --- | --- | --- | --- | --- | --- | --- |
| steps_total | steps_total | 1 | 0.170 | 0.088 | 0.054 | 0.189 |
| steps_total | social_usage | 1 | 0.086 | 0.078 | 0.272 | 0.106 |
| steps_total | nonsocial_usage | 1 | 0.166 | 0.104 | 0.111 | 0.285 |
| steps_total | valence | 1 | 0.182 | 0.124 | 0.142 | 0.442 |
| social_usage | steps_total | 1 | 0.423 | 0.183 | 0.020 | 0.651 |
| social_usage | social_usage | 1 | 0.102 | 0.126 | 0.420 | 0.301 |
| social_usage | nonsocial_usage | 1 | -0.047 | 0.102 | 0.645 | 0.236 |
| social_usage | valence | 1 | -0.093 | 0.143 | 0.516 | 0.409 |
| nonsocial_usage | steps_total | 1 | 0.121 | 0.109 | 0.267 | 0.205 |
| nonsocial_usage | social_usage | 1 | 0.077 | 0.167 | 0.645 | 0.420 |
| nonsocial_usage | nonsocial_usage | 1 | 0.500 | 0.120 | 0.000 | 0.229 |
| nonsocial_usage | valence | 1 | -0.013 | 0.205 | 0.948 | 0.642 |
| valence | steps_total | 1 | -0.015 | 0.090 | 0.869 | 0.195 |
| valence | social_usage | 1 | 0.056 | 0.081 | 0.490 | 0.120 |
| valence | nonsocial_usage | 1 | 0.137 | 0.094 | 0.145 | 0.224 |
| valence | valence | 1 | 0.246 | 0.120 | 0.040 | 0.341 |

Between-users network:

| variable 1 | variable 2 | *p* 1->2 | *p* 1<-2 | pcor | cor |
| --- | --- | --- | --- | --- | --- |
| social_usage | steps_total | 0.000 | 0.000 | 0.658 | 0.699 |
| nonsocial_usage | steps_total | 0.060 | 0.110 | 0.233 | 0.348 |
| nonsocial_usage | social_usage | 0.916 | 0.095 | 0.090 | 0.318 |
| valence | steps_total | 0.006 | 0.852 | 0.162 | 0.039 |
| valence | social_usage | 0.521 | 0.742 | -0.029 | -0.024 |
| valence | nonsocial_usage | 0.048 | 0.003 | -0.400 | -0.366 |

Within-user contemporaneous network (estimated post-hoc):

| variable 1 | variable 2 | *p* 1->2 | *p* 1<-2 | pcor | cor |
| --- | --- | --- | --- | --- | --- |
| social_usage | steps_total | 0.106 | 0.138 | 0.155 | 0.101 |
| nonsocial_usage | steps_total | 0.009 | 0.024 | -0.227 | -0.189 |
| nonsocial_usage | social_usage | 0.001 | 0.037 | 0.262 | 0.233 |
| valence | steps_total | 0.391 | 0.600 | 0.068 | 0.035 |
| valence | social_usage | 0.791 | 0.671 | -0.050 | -0.008 |
| valence | nonsocial_usage | 0.200 | 0.188 | 0.148 | 0.130 |

**Table S8**. **Summary of pre-lockdown network analysis relating daily physical activity to smartphone use and self-reported mood.** *N*=22 participants and 36 time points (days) were included in the model. The temporal network included a 1-timepoint (day) lag for predictive relationships. *steps_total*, total daily step count; *social_usage*, total time spent using social media apps; *nonsocial_usage*, total time spent using any other app, per day; *valence,* mean valence of emotions recorded each day (mood); *SE*, standard error; *SD*, standard deviation; *pcor*, partial correlation*; cor*, correlation.

| Number of times temporal effects were included | | | |  |
| --- | --- | --- | --- | --- |
|  | steps_total | social_usage | nonsocial_usage | valence |
| steps_total | **716** | 10 | 103 | 27 |
| social_usage | 104 | **972** | 5 | 24 |
| nonsocial_usage | 36 | 15 | **999** | 28 |
| valence | 34 | 115 | 50 | 24 |
| Number of times between-subjects effects were included | | | |  |
|  | steps_total | social_usage | nonsocial_usage | valence |
| steps_total | - |  |  |  |
| social_usage | 340 | - |  |  |
| nonsocial_usage | 431 | 290 | - |  |
| valence | 595 | 61 | 622 | - |
| Number of times contemporaneous effects were included | | | |  |
|  | steps_total | social_usage | nonsocial_usage | valence |
| steps_total | - |  |  |  |
| social_usage | 0 | - |  |  |
| nonsocial_usage | 31 | **437** | - |  |
| valence | 127 | 0 | 0 | - |

**Table S9. Stability of pre-lockdown network analysis relating daily physical activity to smartphone use**. Values represent number of times (out of 999 successfully estimated models) each network connection was included in the 25% case-drop bootstrap models. Bold type face indicates effects which were included in the original analysis.

| Number of times temporal effects were included | | | |  |
| --- | --- | --- | --- | --- |
|  | steps_total | social_usage | nonsocial_usage | valence |
| steps_total | 322 | 39 | 152 | 135 |
| social_usage | **368** | 149 | 3 | 135 |
| nonsocial_usage | 29 | 60 | **787** | 46 |
| valence | 9 | 24 | 70 | **319** |
| Number of times between-subjects effects were included | | | |  |
|  | steps_total | social_usage | nonsocial_usage | valence |
| steps_total | - |  |  |  |
| social_usage | **786** | - |  |  |
| nonsocial_usage | 365 | 215 | - |  |
| valence | 519 | 119 | **648** | - |
| Number of times contemporaneous effects were included | | | |  |
|  | steps_total | social_usage | nonsocial_usage | valence |
| steps_total | - |  |  |  |
| social_usage | 229 | - |  |  |
| nonsocial_usage | **529** | **568** | - |  |
| valence | 43 | 40 | 5 | - |

**Table S10. Stability of post-lockdown network analysis relating daily physical activity to smartphone use**. Values represent number of times (out of 788 successfully estimated models) each network connection was included in the 25% case-drop bootstrap models. Bold type face indicates effects which were included in the original analysis.

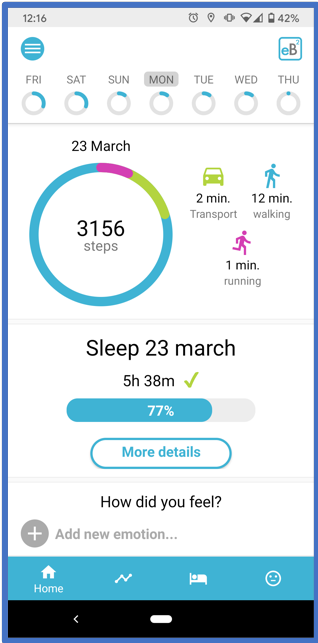

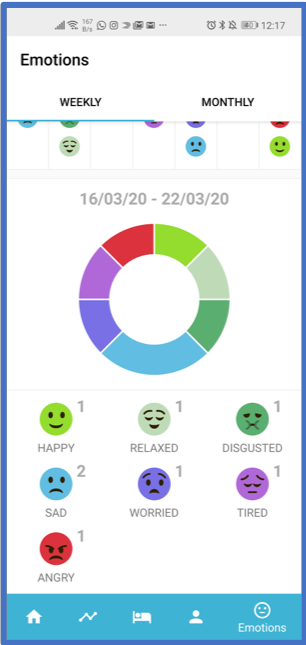

**Figure S1. Representative screenshots from the eB^2^ app.** Left, screen showing daily activity summary data available to the user. Right, interface for ecological momentary assessment of emotional state within the app. Participants were able to open the app and enter information about their current emotional state at any point (as frequently or infrequently as they wanted), by selecting the relevant face icon (emoji). In the version of the app used for data reported here, the available emotions were happy, delighted, motivated, relaxed, neutral, angry, sad, fearful, disgusted, tired, in pain, and worried/overwhelmed. NB, for this study, all app text was in Spanish, therefore English terms represent approximate translations (Spanish language labels were *feliz, encantado, motivado, relajado, neutro, ira, tristeza, asustado, disgustado, cansado, dolor, agobiado*).

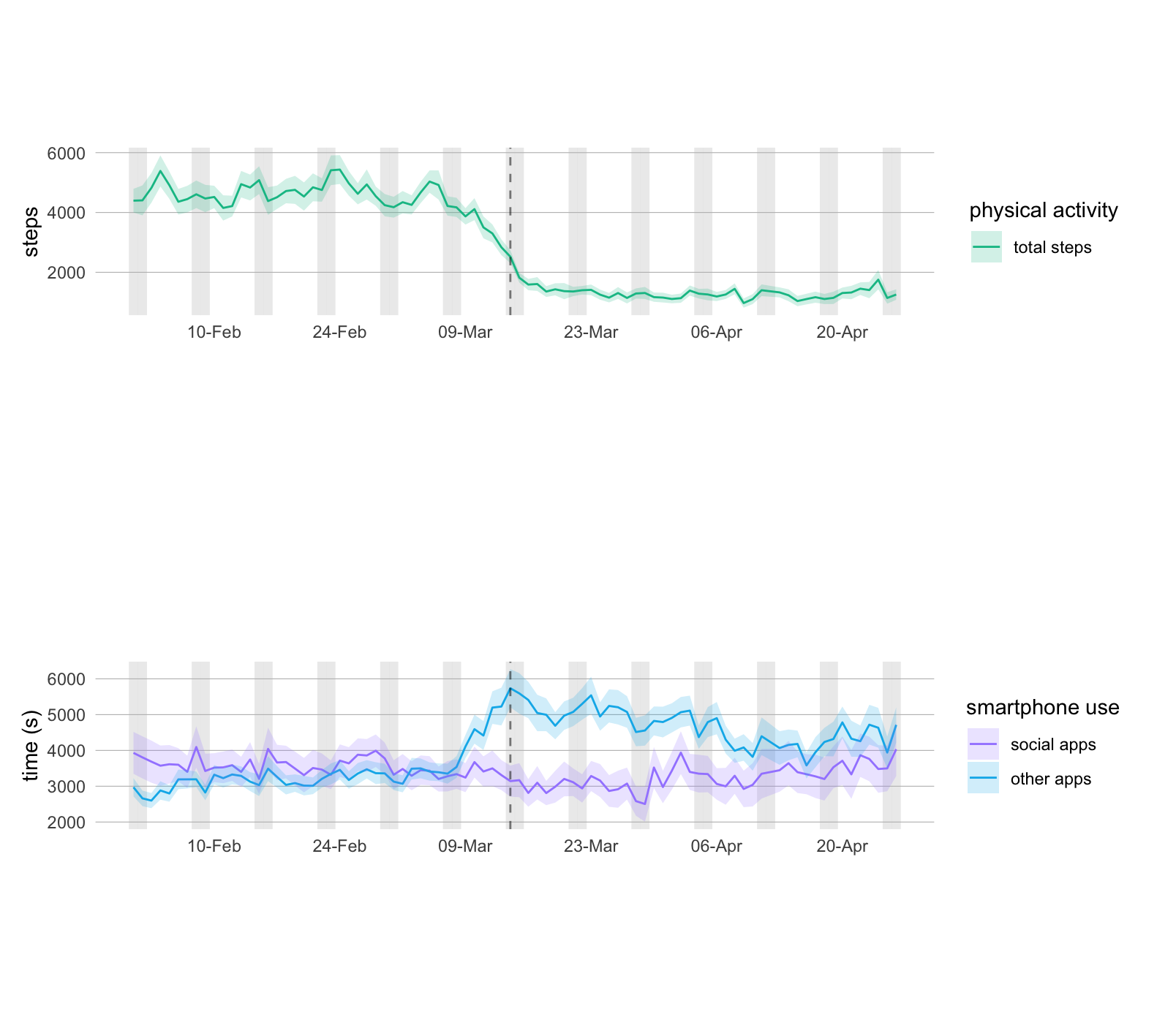

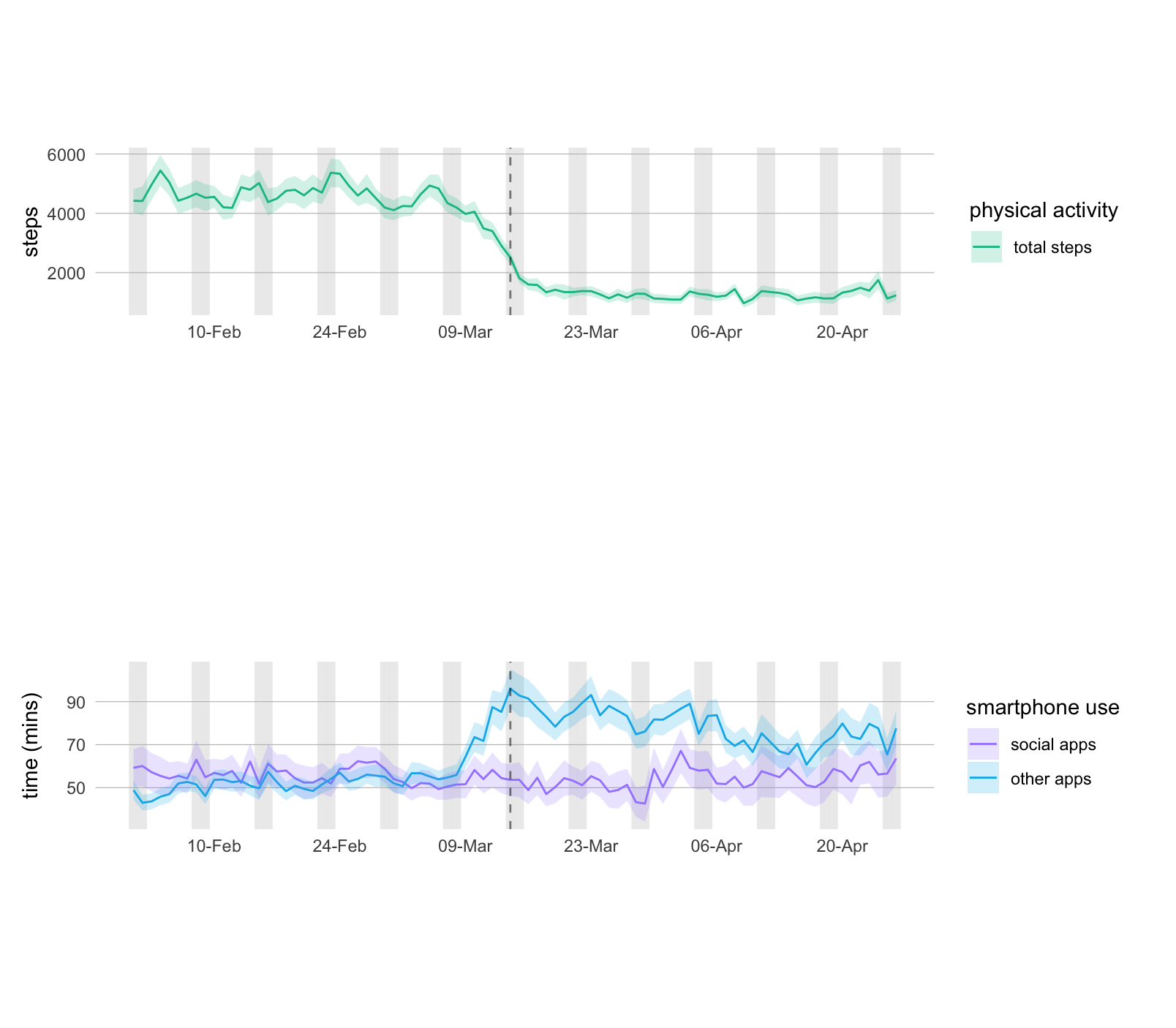

**Figure S2. Physical activity and smartphone use data, following adjustment for weekly seasonality.**

Data represent mean (SE) of daily physical activity (step count), social, and non-social app use, following removal of the 7-day seasonality time series component. The vertical dotted line represents the declaration of a national emergency (and associated lockdown measures) in Spain on 14/03/20. Vertical shading represents weekends (Saturday and Sunday).

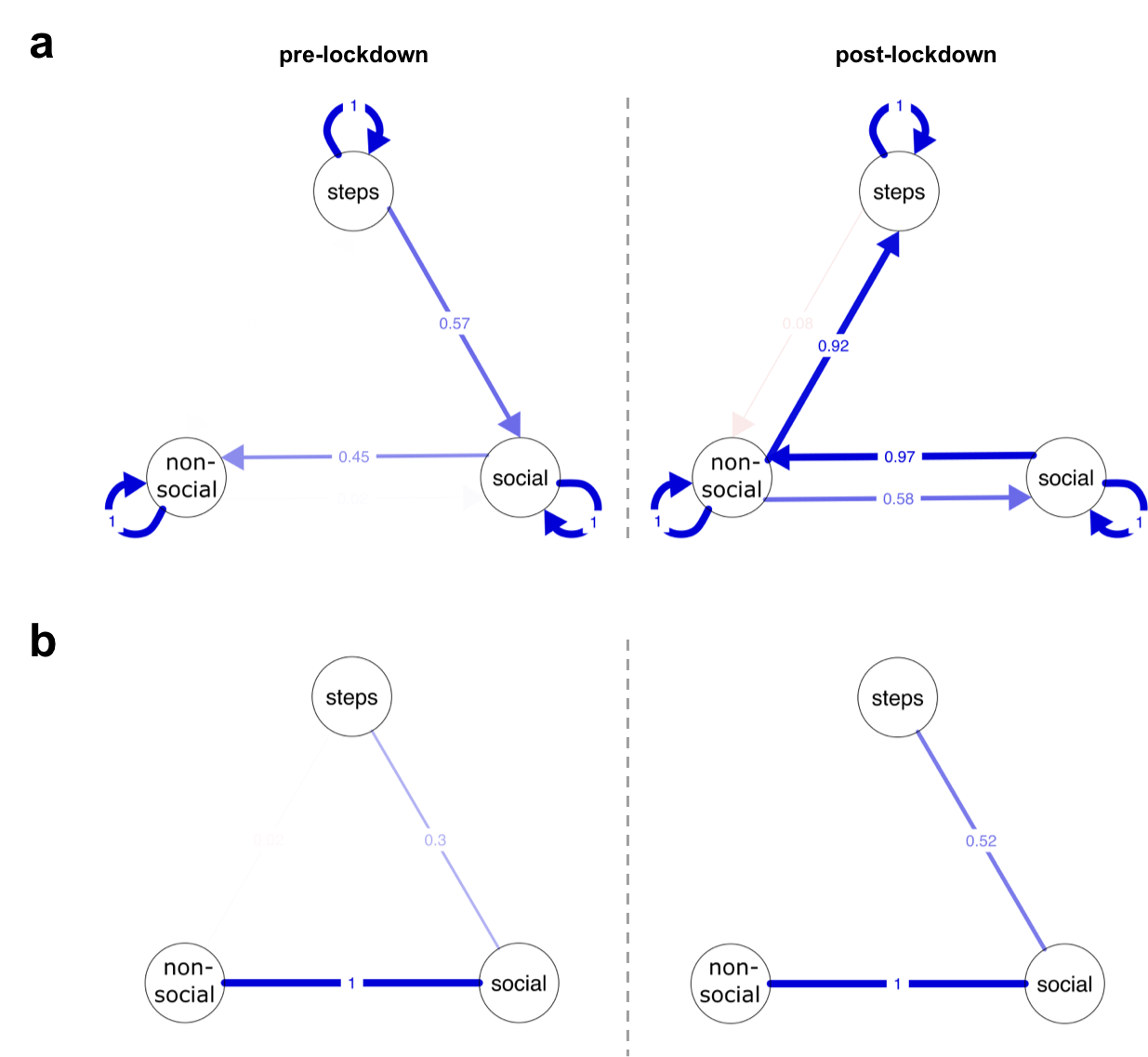

**Figure S3. Results of the case-drop bootstrap model for the physical activity and smartphone use networks**. Edges represent proportion of times each network connection was included in 1000 25% case-drop bootstraps (with the same sign as in the original network), pre- and post-lockdown. **a** For the within-users temporal networks. **b** For the between-users networks. Positive edges are drawn in blue, and negative edges are drawn in red. Edge width is drawn proportional to rate of inclusion across bootstraps.

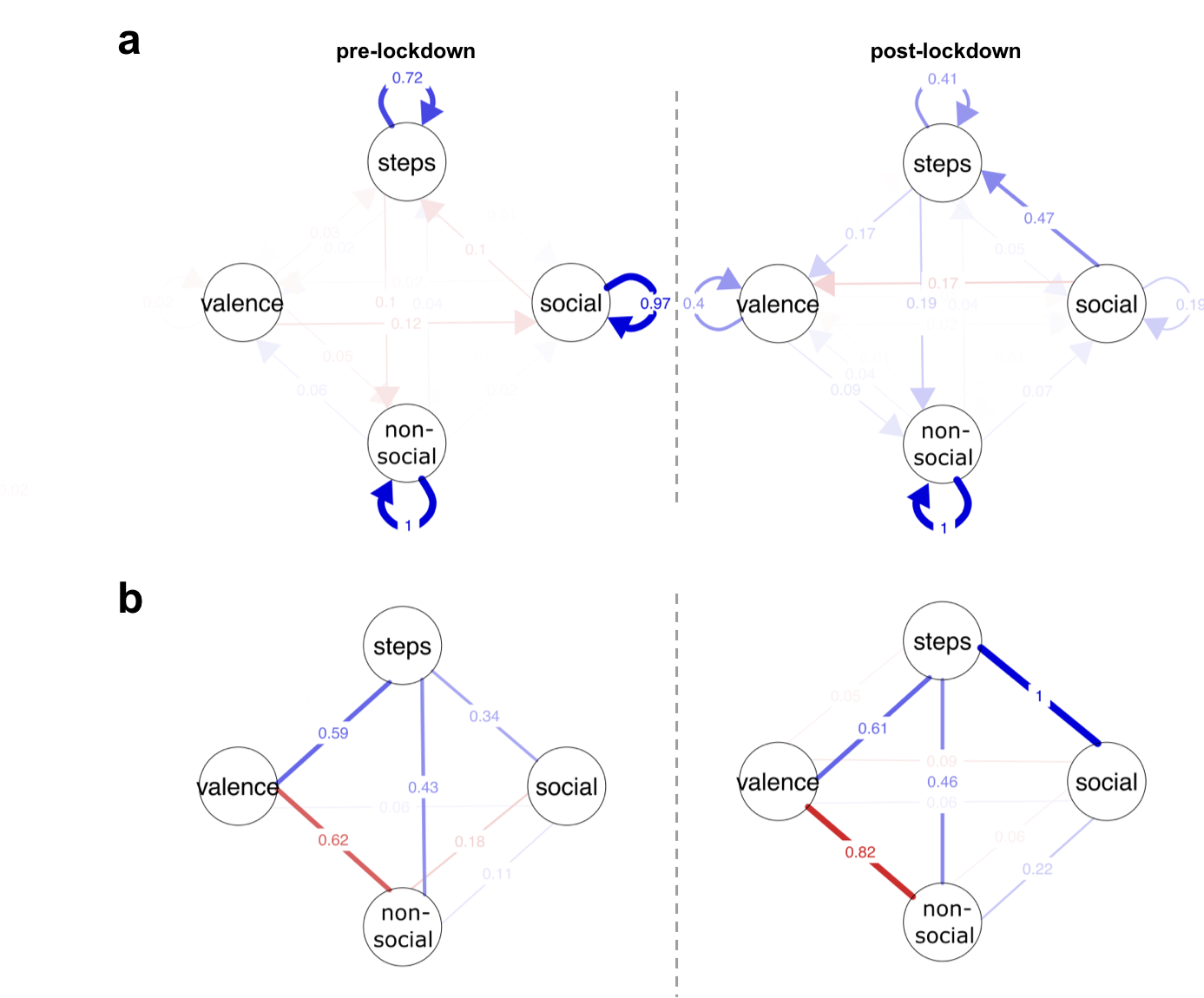

**Figure S4.** **Results of the case-drop bootstrap model for the physical activity, smartphone use, and mood networks**. Edges represent proportion of times each network connection was included in 1000 25% case-drop bootstraps (with the same sign as in the original network), pre- and post-lockdown (for 999 and 788 successfully estimated models, respectively). **a** For the within-users temporal networks. **b** For the between-users networks. Positive edges are drawn in blue, and negative edges are drawn in red. Edge width is drawn proportional to rate of inclusion across bootstraps.
